# MRI-T2 Relaxometry is Increased in Mild Traumatic Brain Injury: Indications of Acute Brain Abnormalities after Injury

**DOI:** 10.1101/2024.05.23.24307520

**Authors:** Mayan J. Bedggood, Christi A. Essex, Alice Theadom, Helen Murray, Patria Hume, Samantha J. Holdsworth, Richard L.M. Faull, Mangor Pedersen

**Author notes:** **Funding:** This project was funded by a grant from the Health Research Council of New Zealand (HRC), grant #21/622.

## Abstract

**Intro:** Mild traumatic brain injury (mTBI) is a common condition, particularly pervasive in contact sports environments. A range of symptoms can accompany this type of injury and negatively impact people’s lives. As mTBI diagnosis and recovery largely rely on subjective reports, more objective injury markers are needed.

**Methods:** The current study compared structural brain MRI-T2 relaxometry between a group of 40 male athletes with mTBI within 14 days of injury and 40 age-matched male controls.

**Results:** Voxel-averaged T2 relaxometry within the grey matter was increased for the mTBI group compared to controls (*p* < 0.001), with statistically increased T2 relaxometry particularly in superior cortical regions.

**Conclusion:** Our findings indicate subtle brain abnormalities can be identified in acute mTBI using MRI-T2 relaxometry. These brain abnormalities may reflect inflammation present in the brain and could constitute an objective injury marker to supplement current subjective methods that dominate clinical decisions regarding diagnosis and prognosis. Future research should validate this potential marker with other data types, such as blood biomarkers or histological samples.

**Significance Statement:** Current understandings of brain pathology underlying mild traumatic brain injury (mTBI) has many gaps and recovery is variable and largely based on subjective reports. Objective markers of injury are required to enhance diagnostic and prognostic capabilities and improve recovery trajectories. Our findings suggest that quantitative MRI-T2 relaxometry times are increased acutely following mTBI compared to controls, possibly indicative of neuroinflammatory processes post-injury. MRI-T2 relaxometry could represent an objective injury marker acutely following mTBI and be utilized to supplement clinical decision making. Insight into mTBI neuropathology could lead to safer, more efficient return to sport, work or education.

## Introduction

Mild traumatic brain injury (mTBI) is a common and disabling brain injury that occurs due to a force to the head caused by a sudden impact, rotational force, rapid deceleration or acceleration (Danielli et al., 2023; McAllister, 2011; Oris et al., 2023; Verboon et al., 2021). This force leads to brain movement within the skull and subsequent focal and diffuse damage (Alam et al., 2020; Verboon et al., 2021). Common symptoms of mTBIs include headaches, difficulty concentrating, sleep difficulties, disturbed balance, confusion, slowed reaction times, nausea and sensitivity to light or noise (Danielli et al., 2023; Di Battista et al., 2020; Markovic et al., 2021; Verboon et al., 2021). Therefore, mTBI can impact patients’ lives in diverse ways and can be associated with reduced work productivity and increased risk of psychiatric and neurodegenerative diseases (Theadom et al., 2017; Wang et al., 2021). Referring to an mTBI as ‘mild’ may not correspond to the personal experience of many people, as many suffer enduring symptoms that persist beyond the expected recovery time and can burden the individual for weeks, months or even years (Danielli et al., 2023; Kara et al., 2020; Rosen, 2024; Slavoaca et al., 2020).

The primary injury in mTBI is from the immediate mechanical damage to the brain that can result in a combination of focal coup and contrecoup damage as well as diffuse damage (Alam et al., 2020; Toma & Nguyen, 2020). Following this, a cascade of secondary injuries can occur, in which inflammation is a key component (Markovic et al., 2021; Piao et al., 2013; Rosen, 2024). Neuroinflammation is a dynamic response of the brain’s immune system to harmful stimuli or events and is initiated by the activation and recruitment of immune cells by facilitating the production of cytokines and chemokines (Pasternak et al., 2012; Pasternak et al., 2016). Microglia play a crucial role in responding to inflammatory events as they survey the brain, phagocytose pathogens and cellular debris, and release inflammatory mediators to aid in the clearance of necrotic tissue to prevent further spread of the injury (Alam et al., 2020; Pasternak et al., 2012; Simon et al., 2017; Strogulski et al., 2023; Yu et al., 2022).

Neuroinflammation can be beneficial in the acute phase following brain injury, promoting repair of damaged tissue, possibly inducing neurogenesis and reducing risk of infection (Monsour et al., 2022). However, when inflammation occurs in excess, it can contribute to loss of neurons and death of brain tissue, which, in some cases, represents a preventable secondary injury cascade (Corps et al., 2015; da Luz Scheffer & Latini, 2020; Kim et al., 2023; Markovic et al., 2021; Mee-Inta et al., 2019; Pasternak et al., 2016).

In the brain, free water is found within the ventricles as cerebrospinal fluid and surrounding the brain parenchyma. However, free water can also accumulate in the extracellular space of the parenchyma due to brain trauma or hemorrhage, for example, that rupture the blood brain barrier (Pasternak et al., 2009). As a result of the inflammatory response following mTBI, a higher amount of water can accumulate in the extracellular space, increasing the free water to bound water ratio. This accumulated fluid, or free water, reflects the fractional volume of unrestricted water and is hypothesized to indicate extracellular changes driven by underlying pathologies (Lyall et al., 2017). This can be a result of multiple different aspects of the inflammatory response, such as inflammatory edema or microglia and other immunoreactive cells (e.g. astrocytes) that induce excessive osmosis of water into the extracellular space from the blood-brain barrier (Albrecht et al., 2016; Lyall et al., 2017; Pasternak et al., 2012).

MRI-T2 relaxometry is a measure of the loss of spin coherence of excited water molecules and subsequently represents the dynamic nature of water and its interaction with macromolecules in tissue (Albrecht et al., 2016). Water bound with larger macromolecules has a spin frequency comparable to the Larmour frequency and is linked with shorter T2 relaxation times. In contrast, free water molecules are smaller, with a faster spin frequency and a longer T2 relaxation time (Cheng et al., 2012; Ghugre et al., 2011; Liu et al., 2018). In healthy tissue, these types of water exist in equilibrium. However, bound water is released in some pathological conditions, increasing the free-water molecule ratio. This creates a medium inefficient for T2 relaxation, subsequently increasing T2 relaxation time as the free water content increases (Cheng et al., 2012; Ghugre et al., 2011; Liu et al., 2018). As T2 relaxometry assesses microstructural tissue alterations, primarily based on water content, it is possible that it may provide a marker of cellular injury and related inflammatory processes (Adel et al, 2023; Cheng et al., 2012; Ghugre et al., 2011; Gracien et al., 2016; Liu et al., 2018; Maillard et al, 2004; Pell et al., 2004; Winston et al., 2017; Yang et al., 2015). For mTBI patients, increased T2 relaxometry times may reflect fluid accumulation in the brain as a result of an increase in the ratio of free water to bound water, due to inflammatory processes characteristic of the secondary injury cascades associated with mTBI.

To the author’s knowledge, only two previous case series have used this technique in mTBI. Pedersen et al. (2020) used MRI-T2 relaxometry and identified potential markers of brain inflammation in a professional Australian Rules football player who had incurred multiple mTBIs. The abnormally elevated T2 relaxometry persisted throughout each symptomatic MRI scan and normalized as the patient recovered. However, T2 relaxation times remained elevated compared to baseline on the last ‘recovery’ scan, suggesting that there may also have been a chronic increase of T2 relaxometry after repetitive mTBIs. In addition, our previous case series analysis (Bedggood et al., 2024) compared T2 relaxometry for 20 individual mTBI participants with the average T2 relaxometry for 44 healthy controls. T2 relaxometry was increased in 19/20 (95%) mTBI participants in at least one cluster of voxels. In five mTBI participants, the areas of increased T2 relaxometry were, at least, partially resolved on recovery re-scans. The aforementioned research demonstrates the potential utility of this MRI method in deepening our understanding of brain pathology following acute mTBI, while highlighting a need for larger group analyses.

Objective markers of brain injury are required to improve mTBI diagnosis and aid in recovery predictions. Given that hospital-based neuroimaging post-mTBI often shows no findings (Mayer et al., 2015), objective measures such as T2 relaxometry may enable clinical decision support with additional objective evidence, complementing subjective self-reporting and clinical symptomatology. Our previous case series (Bedggood et al., 2024) assessed the potential for MRI-T2 relaxometry to identify and quantify subtle brain abnormalities on an individual level. Our current study extends this research by conducting statistical analysis on a group level with a substantially larger mTBI cohort. With this, we increase the power and external validity of the findings and contribute to a significant gap in existing research with regards to MRI-T2 relaxometry and brain trauma. For the current study, we hypothesized that the mTBI group would have significantly increased brain T2 relaxometry compared to the control group, potentially indicating acute neuroinflammation following mTBI.

## Materials and Methods

Ethics approval was obtained from the Health and Disability Ethics Committee, New Zealand (HDEC – 2022 EXP 11078). Institutional approval was also obtained (AUTEC reference 22/12).

### Participants

We recruited two groups between 16 and 35 years of age. The first group consisted of 40 males with a mean age of 21.4 years (± 5.0 years) with acute sports-related mTBI (≤ 14 days post-injury). Participants for this group were included if their injury occurred in a sports game or training and was diagnosed by a doctor. The second group consisted of 40 healthy male controls (athletes and non-athletes) with a mean age of 22.1 years (± 3.8 years) who had not suffered an mTBI in the last 12 months, had not suffered more than five total mTBI in their lifetime and did not have lingering symptoms from a previous mTBI (to minimize potential confounding effects of cumulative damage and/or chronic inflammation). Participants for either group were excluded if they had an existing significant neurological condition or were contraindicated for an MRI scan (e.g., certain metal implants, or braces). An independent-sample t-test showed no significant age difference between the groups, *t*(78) = 0.71, *p =* 0.48.

mTBI participants were recruited at the Axis Sports Concussion Clinics in Auckland, New Zealand and via community links (e.g. physiotherapists, word of mouth, digital and print advertisements). Participants in the mTBI group comprised 27 rugby players, 5 football players, 2 hockey players, 2 martial artists, 1 futsal player, 1 gymnast, 1 surfer and 1 swimmer. Participants were scanned, on average, 10.7 ± 2.9 days following their injury. Control participants were recruited via print advertisements at the Auckland University of Technology and The University of Auckland and social media advertisements. All participants were provided with study information prior to providing informed consent. See Supplementary Materials for additional information on mTBI participants.

### Magnetic Resonance Imaging Procedure

All magnetic resonance images were acquired using a 3T Siemens MAGNETOM Vida fit scanner (Siemens Healthcare, Erlangen, Germany) located at the Centre for Advanced Magnetic Resonance Imaging (CAMRI) at The University of Auckland, New Zealand, using a 20-channel head coil. A T2 mapping sequence was collected to investigate anatomical T2 relaxometry. T2 maps were acquired using an 8 echo Carr-Purcell-Meiboom-Gill (CPMG) sequence (TEs = 28.9, 57.8, 86.7, 115.6, 144.5, 173.4, 202.3 and 231.2 ms; TR = 6s; slice thickness = 2.0mm; voxel size = 2.0 × 2.0 × 2.0mm; matrix size = 112 × 128 × 63; flip angle (FA) = 180°; base resolution = 128; phase resolution = 100%; phase field of view (FOV) = 87.5%). Total T2 mapping acquisition time was 12:02 min. T1-weighted anatomical images were collected for quality control purposes. T1 weighted images were acquired using a magnetization-prepared rapid gradient echo (MPRAGE) sequence (TR = 1.9s; TE = 2.5ms; TI = 979ms; FA = 9°; slice thickness = 0.9mm; voxel size = 0.4 × 0.4 × 0.9 mm; matrix size = 192 × 512 × 512; phase FOV = 100%). Total T1-weighted acquisition time was 4:31 min. A radiologist reviewed clinically relevant MRI images from each participant to check for clinically significant abnormalities that might require further attention.

### Data Processing and Statistical Analysis

All MRI images were received in *DICOM* format, converted to *NIfTI* format, and arranged according to the *Brain Imaging Data Structure (BIDS)* (Boré et al., 2023). Image quality assurance checking was conducted in the *MR View* toolbox of *MRtrix*3 (Tournier et al., 2019) by two investigators in the study (MJB, MP) using the participants’ T1 weighted image as the underlay and their T2 map as the overlay to check for artifacts or abnormalities caused by scanning or processing. Then, preprocessing was conducted on the T2 maps using *PyCharm* version 2022.2.3. This preprocessing step included brain extraction (skull stripping) using the *bet* function in FSL (Smith, 2002) before normalizing each image to the MNI standard space by registering them to a MNI152 2.0 × 2.0 × 2.0mm template image using *FSL FLIRT* (Jenkinson et al., 2002). The third T2 echo volume (86.7 ms) was extracted for each participant to generate a group average brain image and binary grey matter, white matter, and cerebrospinal fluid masks. Lastly, we removed the first volume, using the offset as a fitting parameter to the T2 relaxation data (Milford et al., 2015) and a monoexponential function at each voxel was fitted across all eight echo-times using *qMRLAb* (Karakuzu et al., 2020) in *Matlab R2022B* to calculate the T2 relaxation time for each participant. Note, MRI acquisition and processing details up until this point are the same as specified in our previous case series (Bedggood et al., 2024).

An in-house script in Matlab was then used to convert files back to *NIfTI* format and apply a smoothing of 6mm FWHM. Each participant’s average brain T2 relaxation was then calculated by removing T2-relaxometry voxels with >160ms (i.e. artifactual relaxation times) and applying a grey matter mask to all voxels. Using the *DPABI Statistics Tool* (Yan et al., 2016), a two-sample t-test was conducted with a two-tailed hypothesis. A second grey matter mask was included at this stage to ensure that any voxels smoothed beyond the grey matter were excluded from the final statistical analysis. An independent-sample t-test was conducted in *Jamovi* version 2.3.19.0 (The Jamovi Project, 2024) to quantify whole-brain averaged differences in T2 relaxometry between mTBI and controls. Voxel-wise permutation testing with Threshold Free Cluster Enhancement (TFCE) was conducted in the *DPABI Toolbox* (Yan et al., 2016) with 20,000 permutations to reduce the type 1 error rate and it is relatively insensitive to varying smoothing parameters (Smith & Nochols, 2009).

## Results

### Analysis 1: Average Whole-Brain T2 relaxometry between mTBI and Controls

A two-tailed independent t-test was conducted to determine if there was a difference in mean total grey matter T2 relaxometry between the mTBI and control groups. The mTBI group (70.3 ± 1.55 ms) had significantly higher average total brain T2 relaxometry compared to the control group (69.1 ± 1.48 ms), *t*(78) = 3.56, *p* < 0.001 (see Figure 1). To ensure that total intracranial volume (TIV) did not have an impact on group differences in T2 relaxometry, we also conducted an independent samples t-test on TIV between mTBI and controls, which was not significant, *t*(78) = 1.25, *p* = 0.216.

**Figure 1.**
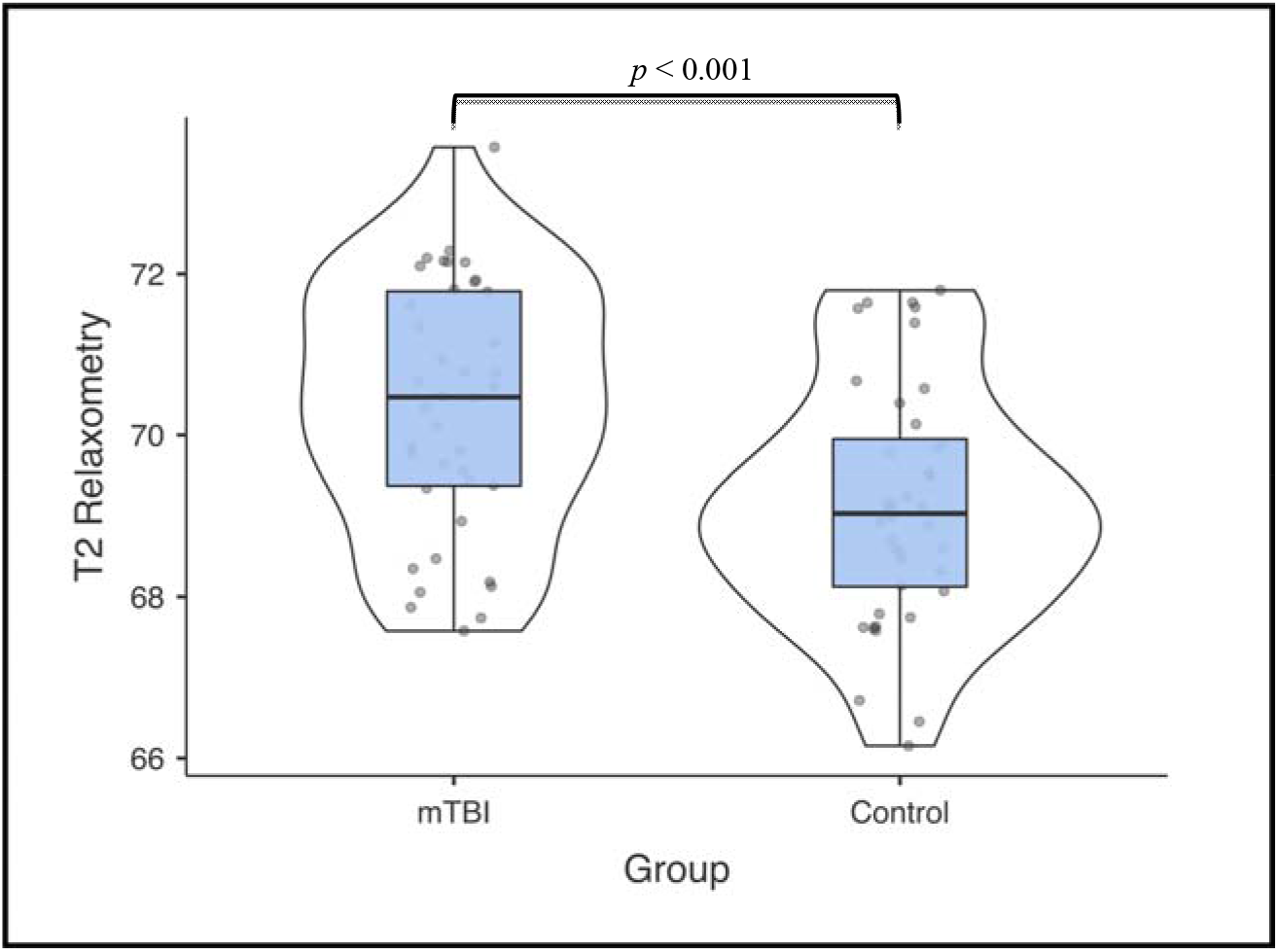
Violin plot depicting statistically significant group difference in average whole-brain T2 relaxometry for mTBI versus controls. An independent sample t-test was used to calculate the difference between groups (using Jamovi), with a significance threshold of *p* < 0.05. Average T2 relaxation time is indicated by the y-axis and measured in milliseconds (*ms*). The median is indicated by a solid center line for each group.

### Analysis 2: Significant Voxels Compared between mTBI and Controls

There were significant differences between mTBI and controls, using a TFCE permutation testing correction for multiple comparisons. These differences were driven by elevated T2 relaxometry in dorsal cortical brain regions of mTBI participants, with fewer statistical differences observed in the inferior regions. Elevated T2 relaxation times in the mTBI group were observed in multiple anatomical regions, including the superior frontal gyrus, middle frontal gyrus, inferior frontal gyrus, cingulate gyrus, dorsolateral prefrontal cortex (DLPFC), anterior cingulate cortex, superior parietal lobe, precentral gyrus, postcentral gyrus, inferior parietal lobe, insula, middle temporal gyrus, superior temporal gyrus, temporoparietal junction (TPJ), and cuneus of the occipital lobe (see Figure 2).

**Figure 2.**
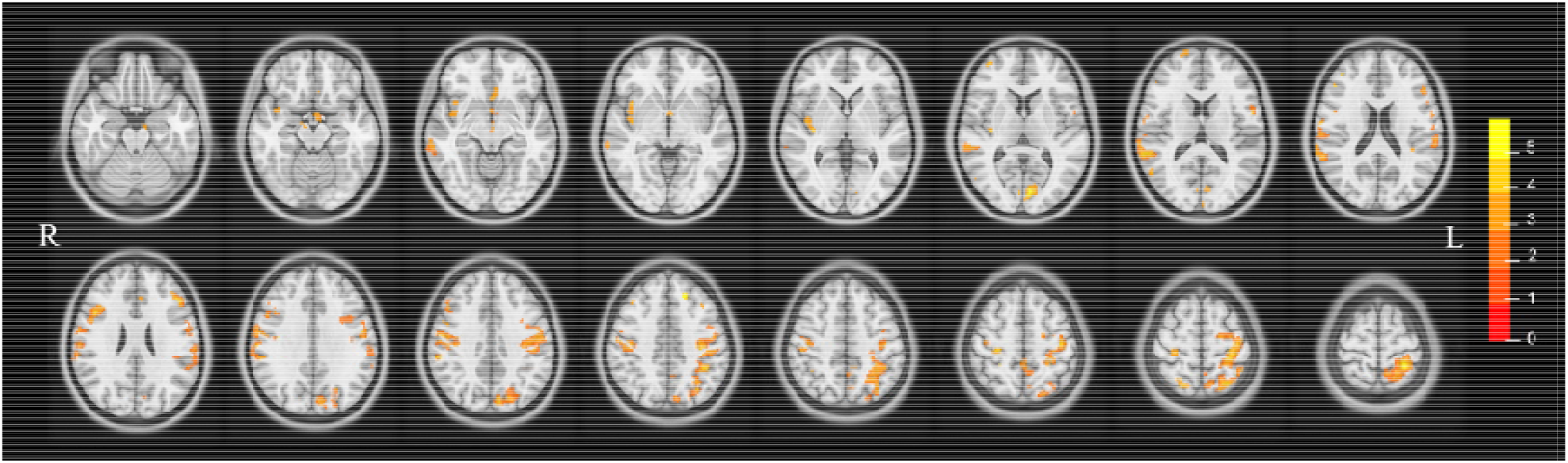
The above figure illustrates group differences in average T2 relaxometry across the whole-brain, measured in t-values. This finding indicates significantly increased T2 relaxometry in the mTBI group compared to the control group.

The commonly used Yeo 7 network parcellation mask (Yeo et al., 2011) was also used to quantify the proportion of TFCE voxels within each of the 7 networks in the brain (see Figure 3) that were significantly different between mTBI and controls. The highest proportion of voxels that were significantly increased for mTBI compared to controls were significant in cortical networks, including the somatomotor network (11% of voxels), dorsal attention network (9% of voxels), and ventral attention areas (7% of voxels). Less significant voxels were seen in the fronto-parietal network (4% of voxels), visual network (2% of voxels), default mode network (1% of voxels), and limbic network (0.5% of voxels).

**Figure 3.**
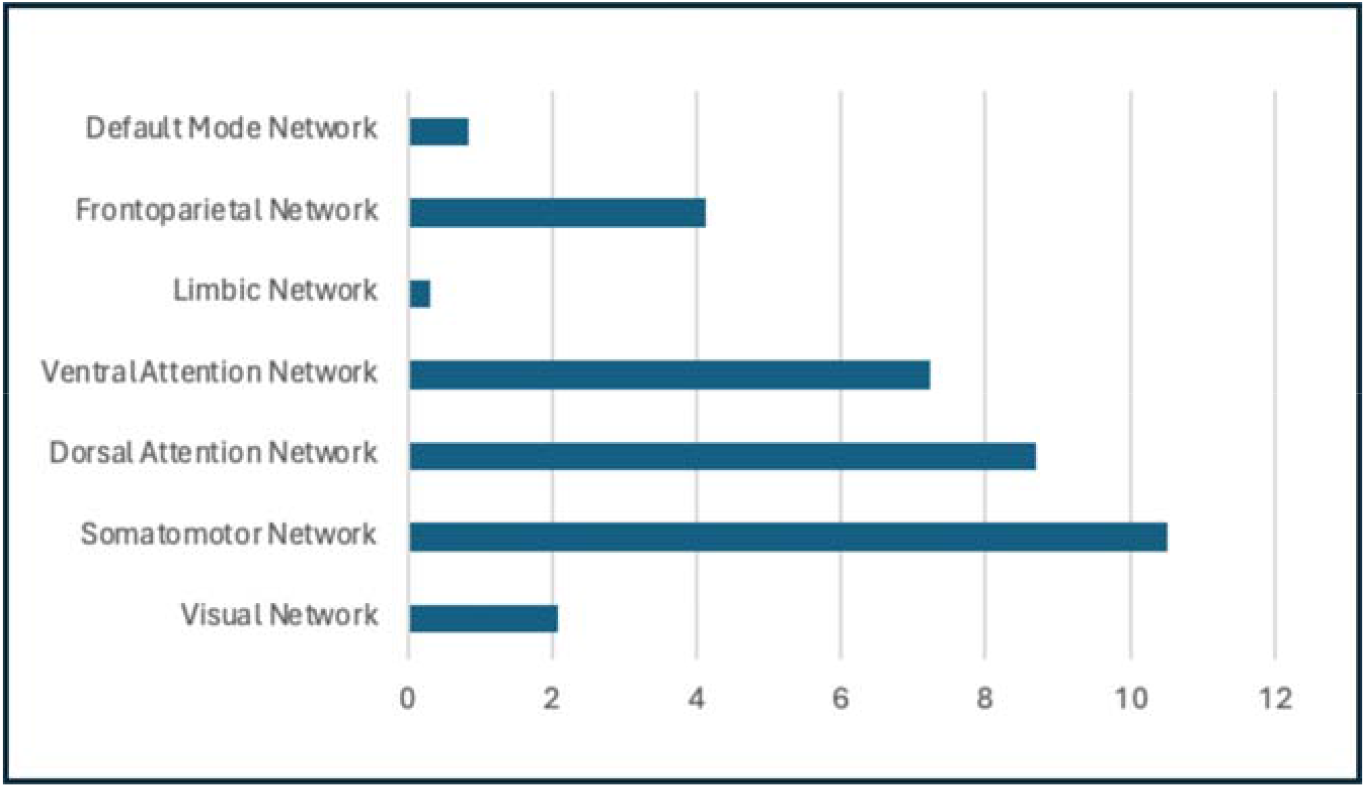
Proportion of Voxels within Brain Networks that are Significantly Increased for mTBI compared to Controls (%).

## Discussion

The current study aimed to investigate the potential of quantitative MRI-T2 relaxometry to be used as an objective marker of injury. We found that brain MRI-T2 relaxometry was increased acutely following sports-related mTBI compared to healthy controls (Figure 1), which supports our study’s hypothesis. T2 relaxation times are influenced by the water content of tissue, with longer relaxation times observed in tissue with a higher ratio of free water molecules compared to bound water molecules. This ratio of water molecules can be observed in conditions including disease and trauma and may be reflective of the brain’s inflammatory response. Inflammation encapsulates many processes and can lead to increases in extracellular free water (Albrecht et al., 2016; Lyall et al., 2017; Pasternak et al., 2012). While we do not know if our findings represent increased intracellular and/or extracellular fluid accumulation, as we cannot directly test this with MRI-T2 methods, we hypothesize that our results are indicative of inflammatory processes acutely post-mTBI. This finding may be due to the inflammatory response that is a part of the secondary injury cascade following mTBI, which aligns with our study timeframe (within 14 days post-injury).

There is existing support for the notion that excessive fluid accumulation could have utility as a biomarker of neuroinflammation (Pasternak et al., 2012). For example, a significant relationship has been found (Lesh et al., 2021) between a known inflammation-related metabolite, glutathione (GSH), and free water, supporting the idea that free water indicates an inflammatory response in the brain. Increased free water has been found in white matter associated with small vessel disease (SVD), which is suggested to indicate neuroinflammation (Duering et al., 2018; Sun et al., 2024). Furthermore, free water has been correlated with elevated levels of brain interferon-gamma, a cytokine that has a crucial role in an array of immune responses, solidifying the sensitivity of free water as a marker of neuroinflammation (Febo et al., 2020; Sun et al., 2024). Finally, further support for the hypothesis that increased water volume represents neuroinflammation comes from schizophrenia research, where abnormalities in the immune response play a key role in the pathophysiology (Lyall et al., 2017; Pasternak et al., 2016). The link between free water and neuroinflammation needs to be validated further, given that free water may be influenced by multiple factors, including neuronal degeneration, hemorrhage or edema. Validation could be done by assessing correlations with more established measures of immune function, such as those using histological samples.

It is important to note, however, that prior research supporting the link between increased water and inflammatory processes is dominated by a type of diffusion MRI method. While both diffusion and T2 relaxometry are sensitive to extracellular water changes, they are measuring different properties. Diffusion techniques may be able to show free water increases during an inflammatory response as inflammation can lead to a breakdown in cellular barriers and subsequently increase the amount of free water that can be detected by diffusion MRI. Therefore, while diffusion techniques more directly measure the mobility of water molecules, T2 relaxometry revolves around water-protein interactions and the tissue microenvironment. While it is useful and relevant to consider research using free water diffusion methods when assessing a potential link to neuroinflammation, caution should be taken with direct comparisons to the potential link between T2 relaxometry and neuroinflammation. Future research could endeavor to utilize both methods and analyse correlations between them.

MRI-T2 relaxometry has previously been utilized in various clinical research settings, including hippocampal sclerosis research (E.g., Adel et al., 2022; Bartlett et al., 2007; Jackson et al., 1993; Winston et al., 2017), juvenile dermatomyositis (E.g., Maillard et al., 2004), multiple sclerosis (E.g., Gracien et al., 2016) and in a rodent model of mTBI (E.g., Yang et al., 2015). However, to the author’s knowledge, only two previous studies have looked at MRI-T2 relaxometry in human mTBI (Bedggood et al., 2024; Pedersen et al., 2020) and the current study is the first research looking at its use, on a group level, in a cohort of mTBI. Our results were consistent with a previous case study that found increased T2 relaxometry in a sports player with an acute mTBI (Pedersen et al., 2020) and our case series (Bedggood et al., 2024) that also demonstrated increased T2 relaxometry in individuals with acute mTBI compared to controls. It is possible that increased T2 relaxometry reflects other processes, such as hemorrhage, changes in iron, molecular changes in the tissue composition or tissue necrosis (Pasternak et al., 2016). However, we did not see evidence of hemorrhage, tissue necrosis would be unlikely in a mild injury like mTBI, and our previous research, while limited, has suggested that T2 relaxometry times may normalize as the patient recovers (Bedggood et al., 2024; Pedersen et al., 2020), which is consistent with a transient inflammatory process post-injury, as opposed to a more permanent loss of neuronal tissue. Therefore, we believe the increased T2 relaxometry in the mTBI group is more likely to be indicative of subtle brain inflammation post-mTBI. However, further research is needed to eliminate other possible causes of increased T2 relaxometry.

We observed that increased T2 relaxometry was widespread but particularly prominent in superior dorsal regions of the frontal and parietal lobes. Specific anatomical regions with elevated T2 relaxation times included areas such as the superior, middle and inferior frontal gyri, the cingulate gyrus, the DLPFC, anterior cingulate cortex, precentral and postcentral gyri, the insula, middle and superior temporal gyri, the TPJ, and cuneus (see Figure 2). Cortical networks with elevated T2 relaxation times included the somatomotor, dorsal attention and ventral attention networks. The dorsal cortical location of increased T2 relaxometry could represent areas of increased vulnerability following mTBI. This could be due to where coup and contrecoup damage commonly occur or where there is likely to be more concentration of force in cortical sulci, for example (Drew & Drew, 2004; Kornguth et al., 2017; Toma & Nguyen, 2020). Our findings overlap with existing research indicating a predictable profile of brain regions that are vulnerable in mTBI, including the frontal and temporal lobes in general and, more specifically, the dorsolateral prefrontal cortex, orbitofrontal cortex, anterior cingulate cortex, temporal polar cortex, amygdala, hippocampi, cerebellum and brainstem (Bigler, 2023; McAllister, 2011). Although previous research has shown that T2 relaxometry decreased after clinical recovery, it remains unknown what the longer-term implications are, especially as cortical regions are known to be an early marker for cumulative damage and chronic injury, including neurodegenerative disease (Bieniek et al., 2021; Ghajari et al., 2017; McKee et al., 2016; McKee et al., 2018).

One of the multi-faceted challenges that accompany mTBI is that whilst some people recover quickly with no intervention, many experience chronic symptoms and functional deficits. Therefore, there is a need to identify those at risk of slowed recovery so clinicians can provide individualized treatment where necessary. Although FDA-approved blood biomarkers (ubiquitin C-terminal hydrolase-L1 and glial fibrillary acidic protein) are promising from an acute clinical perspective, there are currently no reliable biomarkers of mTBI that could be utilized a week or two after injury, when many patients present to clinics (Diaz-Arrastia et al., 2014; Kobeissy et al., 2024). Furthermore, the process of recovery relies on subjective symptom reporting. Enhancing our understanding of neurological changes that take place post-mTBI may identify those at a greater risk of poor recovery trajectories. A reliable biomarker of inflammation is important as it may provide a time window for anti-inflammatory treatment in the acute stages post-mTBI, which could minimize secondary injury cascades relating to neurodegenerative processes (Pasternak et al., 2012). Ultimately, having an objective marker of injury and/or recovery from mTBI has the potential to contribute to a safer and more efficient return to play, education or employment.

The current study has limitations that should be considered when evaluating the findings. Firstly, we only included male participants, which will reduce external validity and transferability to wider mTBI populations. If both sexes were to be included, separating them into distinct groups and comparing them would be most sensible, as there may be sex differences in presentation following mTBI (e.g., due to differences in hormones at the time of injury or neck musculature). Creating this between-group comparison would require a larger sample size and reduce the feasibility of the study. The selection of only males allows for a substantial sample size to be collected for group comparison of mTBI versus controls to test the feasibility of imaging markers in a homogenous sample. Larger future studies should include females too and look at group differences (e.g., looking at the influence of estrogen and progesterone hormones for females during the acute phase of injury), increasing the generalizability and clinical utility of results. While we made efforts to recruit participants as soon after their injury as possible, the MRI scans varied within a 14-day time frame. Future research should endeavor to scan participants within a shorter time frame to increase the homogeneity between injury and scan, reducing possible timing confounds. In addition, re-scanning all participants upon clinical recovery to see if elevated T2 relaxometry resolves with symptom resolution and whether it could be used to predict recovery, would be beneficial. Doing so could strengthen the link between this objective marker and subjective reports and help decipher the transient versus chronic nature of the inflammatory response instigated by an mTBI.

Further research is needed to support the potential of T2 relaxometry as an imaging marker of mTBI injury. As the increase in extracellular volume within a voxel could be caused by a multitude of processes, we cannot be certain that the increased T2 relaxometry that we found is indeed indicative of inflammation. Validating the MRI-T2 relaxometry findings with blood biomarkers or histological measures of inflammation would be beneficial in strengthening the clinical utility of this potential injury marker. Lastly, future research could supplement the current approach by utilizing free water imaging methods too. While T2 relaxometry can identify changes in water content via molecular interactions, free water imaging can identify extracellular volume more directly. It is possible that this is more specific to neuroinflammation and therefore could strengthen current T2 relaxometry findings (Pasternak et al., 2016).

## Conclusion

MRI-T2 relaxometry was increased in athletes with mTBI compared to healthy controls. Increased T2 relaxometry was widespread but particularly prominent in the dorsal cortices of the brain. Our results suggest possible brain inflammation acutely following mTBI and could represent an objective injury marker for diagnostic and prognostic purposes. Future research should replicate this finding and validate whether advanced MRI methods, alongside other biomarkers, have clinical utility.

## Data Availability

All data produced in the present study are available upon reasonable request to the authors

## Author Contributions

**Mayan J. Bedggood** (Conceptualization, Methodology, Project Administration, Investigation, Validation, Resources, Data Curation, Software, Formal Analysis, Visualization, Writing - Original Draft, Writing - Review & Editing); **Christi A. Essex** (Writing - Review & Editing, Project administration, Investigation); **Alice Theadom** (Funding acquisition, Conceptualization, Methodology, Writing - Review & Editing, Supervision); **Helen Murray** (Writing - Review & Editing); **Patria Hume** (Writing - Review & Editing); **Samantha J. Holdsworth** (Writing - Review & Editing); **Richard L.M. Faull** (Writing - Review & Editing); **Mangor Pedersen** (Funding acquisition, Conceptualization, Methodology, Project Administration, Validation, Resources, Data Curation, Software, Formal Analysis, Visualization, Writing - Review & Editing, Supervision)

## Acknowledgements

Thank you to Amabelle Voice-Powell, Cassandra Mcgregor and Aria Courtney for their contribution to data collection, Tania Ka’ai for her contribution to the concept and design of the study, to Axis Sports Concussion Clinic for their assistance with mTBI recruitment, to the Centre for Advanced Magnetic Resonance Imaging for their assistance with MRI data collection and to Dr Tim Elliot for radiological reporting.

## Data Availability

Data can be made available by request to the corresponding author.

## Supplementary Materials

**Table 1.** Participant Injury and MRI Details.

| **ID** | **Method of Injury** | **Post-injury MRI (days)** | **Prior mTBI** | **BIST Score (/160)** | **Recovery (days)** |
| --- | --- | --- | --- | --- | --- |
| mTBI1 | Rugby | 5 | 0 | 140 | 26 |
| mTBI2 | Rugby | 5 | 1 | 12 | 20 |
| mTBI3 | Rugby | 6 | 0 | 78 | 47 |
| mTBI4 | Rugby | 13 | 4 | 18 | 24 |
| mTBI5 | Rugby | 12 | 0 | 61 | 23 |
| mTBI6 | Rugby | 13 | ? | 42 | 17 |
| mTBI7 | Football | 13 | 1 | 13 | 20 |
| mTBI8 | Hockey | 12 | 1 | 6 | 31 |
| mTBI9 | Rugby | 6 | ? | 56 | 18 |
| mTBI10 | Rugby | 12 | 4 | 54 | 117 |
| mTBI11 | Rugby | 10 | 2 | 52 | 27 |
| mTBI12 | Football | 13 | 0 | 13 | 27 |
| mTBI13 | Rugby | 5 | 2 | 79 | 66 |
| mTBI14 | Rugby | 13 | 0 | 2 | 26 |
| mTBI15 | Rugby | 13 | 1 | 22 | 26 |
| mTBI16 | Futsal | 8 | 0 | 117 | 32 |
| mTBI17 | Rugby | 13 | ? | 0 | 18 |
| mTBI18 | Gymnastics | 10 | 1 | 34 | 56 |
| mTBI19 | Jiu-jitsu | 13 | 2 | 28 | 31 |
| mTBI20 | Surfing | 11 | 5+ | 69 | ? |
| mTBI21 | Hockey | 14 | 1 | 40 | 50 |
| mTBI22 | Rugby | 7 | 1 | 14 | 28 |
| mTBI23 | Rugby | 13 | 6 | 47 | 157 |
| mTBI24 | Football | 12 | 0 | 28 | 146+ |
| mTBI25 | Judo | 14 | ? | 39 | 38 |
| mTBI26 | Rugby | 10 | 2 | 34 | 20 |
| mTBI27 | Rugby | 12 | 3 | 68 | 156 |
| mTBI28 | Rugby | 12 | ? | 17 | ? |
| mTBI29 | Rugby | 12 | 0 | 12 | 19 |
| mTBI30 | Rugby | 12 | ? | 25 | 37+ |
| mTBI31 | Football | 7 | 3 | 30 | 49 |
| mTBI32 | Swimming | 12 | 1 | 51 | 82+ |
| mTBI33 | Rugby | 5 | 5+ | 6 | 12 |
| mTBI34 | Rugby | 12 | 1 | 2 | 19 |
| mTBI35 | Rugby | 14 | 1 | 22 | 21 |
| mTBI36 | Rugby | 12 | ? | ? | ? |
| mTBI37 | Rugby | 12 | ? | 91 | 59 |
| mTBI38 | Rugby | 12 | 0 | 61 | 9 |
| mTBI39 | Football | 8 | 2 | 58 | 78+ |
| mTBI40 | Rugby | 8 | 0 | 8 | 22 |
| **Mean** | NA | 10.65<br>($\pm 2.9$) | N/A | 39.71<br>($\pm 32.0$) | 45.22 |

*Note.* The above table provides additional data for each participant in the mTBI group. Their ‘Method of Injury’ indicates what sport they were engaged in at the time of their mTBI, ‘Post-injury MRI’ is the length of time between their mTBI and MRI scan, ‘Prior mTBI’ is the total amount of previous mTBIs suffered in their lifetime, ‘Total BIST score’ is their overall questionnaire score >24 hours post-injury out of a total of 160, and ‘Recovery’ is the length of time between injury and discharge or recovery as determined by their doctor.

